# Imaging Strategies and Futile Transfers in the Drip-and-Ship Model Within a Densely Connected Stroke Network

**DOI:** 10.64898/2026.05.31.26354563

**Authors:** Po-Yu Tsai, Che-Wei Lin, Yu-Ming Chang, Ray-Chang Tzeng, Ming-Hsiu Wu, Si-Chon Vong, Tsang-Shan Chen, Shang-Te Wu, Yu-Tai Tsai, Yi-Ting Fang, Chuang-Chou Yang, Yu-Hsiang Su, Meng-Hua Huang, Mu-Han Wu, Feng-Yuan Chu, Yen-Chu Huang, Kuan-Hung Lin, Che-Chao Chang, Ching-Han Wu, Chun-Min Wang, Pi-Shan Sung

## Abstract

**Background and Purpose:** Futile interhospital transfers, where patients transferred for endovascular thrombectomy (EVT) do not ultimately receive the procedure, represent a critical systemic burden on stroke transfer network. Whether pre-transfer computed tomography angiography (CTA) at the primary stroke center (PSC) reduces futile transfers, and at what workflow cost, remains incompletely characterized.

**Methods:** This retrospective study enrolled 314 acute ischemic stroke patients transferred for potential EVT within the Tainan-Chiayi Stroke Network (October 2021–September 2025). Patients were stratified by CTA timing: pre-transfer (n=66) versus post-transfer (n=248). Workflow time metrics and 90-day functional outcomes were compared. Futile transfers were classified into three categories: preventable over-triage, physiological futility, and gray zone cases.

**Results:** The futile transfer rate was substantially lower in the pre-transfer CTA group (27.3% vs. 66.1%; P<0.001), with post-transfer CTA as the strongest independent predictor of futility (aOR 5.21; 95% CI 2.83–9.60). In the post-transfer CTA group, 40.2% of futile transfers involved conditions identifiable by pre-transfer CTA. Regardless of CTA timing, gray zone cases predominated in both groups (83.3% vs. 47.6%), driven by intracranial atherosclerotic stenosis/ chronic total occlusion, large infarct cores, and medium vessel occlusions. Pre-transfer CTA significantly prolonged PSC door-in-door-out time (140 vs. 88 min; P<0.001) and showed numerical trends toward longer onset-to-EVT time and lower rates of favorable functional outcome.

**Conclusions:** Adopting CTA during the pre-transfer period reduces preventable futile transfers but prolongs PSC processing time. Nevertheless, the persistent gray zone requires strategies beyond imaging alone, and the trade-off between triage precision and transfer efficiency warrants ongoing evaluation across different stroke networks settings.

## Introduction

Endovascular thrombectomy (EVT) has revolutionized the management of acute ischemic stroke (AIS) due to large-vessel occlusion (LVO)^1^, with randomized evidence demonstrating robust clinical benefit across diverse patient profiles^2–5^. Given that the clinical benefit of EVT is time-dependent^6, 7^, the efficiency of the stroke system of care—encompassing prehospital triage, imaging acquisition, and interhospital workflow—is a critical determinant of patient outcomes.

However, interfacility transfer remains a significant bottleneck. While the time penalty of “drip-and-ship” workflows appears modest in controlled trial settings^8, 9^, it is substantially more pronounced in real-world practice^10^, where logistical and institutional variability amplify transfer-related delays. Compounding this inefficiency, studies across diverse healthcare systems consistently report that 40–60% of patients transferred for potential EVT do not ultimately undergo the procedure^11–13^. These futile transfers consume critical transport resources, occupy comprehensive stroke center (CSC) capacity and expose patients to the risks of interfacility displacement without therapeutic benefit. Reducing this rate has accordingly become a priority for regional stroke network optimization.

Reflecting a global paradigm shift, several guidelines and consensus statements have recently advocated for the routine performance of computed tomography angiography (CTA) at the primary stroke center (PSC) prior to transfer ^14–16^. This strategy aims to transition from symptom-based triage to an image-guided “precision triage” model. By identifying non-LVO cases before transfer, pre-transfer CTA has the potential to substantially reduce unnecessary transfers and ensure that only patients with confirmed vascular targets arrive at the CSC.

However, even among patients transferred with pre-transfer CTA, a substantial gray zone remains. In an era of rapidly evolving EVT indications—where evidence for intervention in large infarct cores^3, 4^, medium vessel occlusions^5^, and atherosclerotic lesions^17, 18^ continues to emerge—the boundary between “appropriate but unsuccessful” and “inappropriate” transfer is inherently fluid and cannot be adjudicated by a single static standard. In East Asian populations, this complexity is further compounded by the high prevalence of intracranial atherosclerotic stenosis (ICAS)^19, 20^, where chronic total occlusions (CTO) or severe stenosis may mimic acute embolic LVO even on CTA, These observations underscore the need for empirical, network-level data to quantify the preventable fraction, characterize the composition of the gray zone, and inform targeted system-level interventions.

Additionally, the applicability of pre-transfer CTA across heterogeneous healthcare environments remains actively debated. The diagnostic benefit of pre-transfer CTA must be weighed against a real but incompletely quantified cost: prolongation of door-in-door-out (DIDO) time at the PSC^21, 22^. In settings lacking 24/7 radiology coverage or on-site technician availability—common features of rural emergency departments and high-volume community centers in both Western and Asian contexts^23–26^—the time required for imaging may substantially delay transfer initiation. This creates a fundamental tension between the “time-is-brain” imperative and the goal of triage precision. Critically, this trade-off has rarely been quantified empirically; most recommendations rest on conceptual arguments rather than network-level transfer efficiency and outcome data.

To address these gaps, we leveraged the Tainan-Chiayi Stroke Network (TCSN)—a regional transfer network covering approximately 2.6 million residents with marked variability in PSC imaging capabilities, operating through a digitally-integrated EVT transfer system (EVTTS)—to provide a granular, system-level analysis^27^. Specifically, this study aimed to: (1) quantify the proportion of futile transfers attributable to preventable causes, and evaluate how pre-transfer CTA modifies this burden; (2) characterize the composition of gray zone futile transfers across both imaging strategies, identifying the dominant underlying causes as a foundation for targeted system improvement; and (3) quantify the workflow phases at which pre-transfer CTA imposes delays and the magnitude of this time cost.

## Methods

### Study Design and Setting

This retrospective observational study was conducted within Tainan and Chiayi Stroke Network (TCSN), a regional healthcare network covering Tainan City, Chiayi City, and Chiayi County in Southern Taiwan. As of October 2025, the demographic data indicates a population of 1,854,000 in Tainan City (density: 845.94 people/km²), 260,584 in Chiayi City (density: 4,341.21 people/km²), and 474,396 in Chiayi County (density: 249.21 people/km²) (Supplementary Figure S1). TCSN comprises 4 CSCs and 11 primary stroke centers (PSCs). Transfer distances from PSCs to CSCs range from 2.6 km (approximately 7 minutes) to 74.5 km (approximately 55 minutes), based on simulated travel times using Google Maps during weekday peak traffic hours.

TCSN operates under a hybrid model combining “Mothership” (MS, direct transport to CSC) and “Drip-and-Ship” (DS; initial intravenous thrombolysis [IVT] at PSC followed by transfer to CSC) strategies. Approximately 23% of patients with LVO in this network require interhospital transfer via the DS model^28^. To enhance transfer efficiency, ensure data security, and facilitate quality monitoring, a web-based EVTTS was implemented in October 2021. Our previous study demonstrated that the implementation of EVTTS significantly improved time metrics and functional outcomes for patients transferred via the DS model^27^, achieving performance comparable to the Mothership model^28^.

### Pre-transfer image strategies in TCSN

Because only 3 PSCs within the TCSN are capable of providing 24-hour CTA services, the TCSN transfer consensus does not mandate CTA or MRA confirmation of LVO before transfer. MRI is generally unavailable at PSCs during the acute pre-transfer stage. In the acute management of suspected stroke, patients initially undergo non-contrast computed tomography (NCCT) to exclude intracranial hemorrhage and determine eligibility for IVT. If LVO is clinically suspected, transfer coordination with a CSC can be initiated via EVTTS even when only NCCT is available. In contrast, PSCs with CTA capability generally perform CTA before transfer and proceed with interhospital transfer only after confirming LVO.

### Study Population

We enrolled patients with acute ischemic stroke who were transferred from PSCs to CSCs via the EVTTS between October 2021 and September 2025. Patients were excluded if they: (1) did not undergo CTA at PSCs or CSCs prior to EVT assessment; (2) had in-hospital stroke onsets or developed LVO symptoms during an emergency department stay at the PSC (to ensure the comparability of time metrics regarding community-onset strokes); (3) was not actually transferred; or (4) had duplicated records.

### Data Collection and Definitions

Baseline demographic and clinical variables were collected, including age, sex, baseline National Institutes of Health Stroke Scale (NIHSS) score, and the administration of reperfusion therapies (intravenous thrombolysis [IVT] and EVT). We extracted detailed time metrics, including onset-to-PSC door time, PSC door-in-to-door-out (DIDO) time, interhospital transfer time, onset-to-CSC door time, and onset-to-CTA time. For process workflow analysis, we calculated PSC/CSC door-to-CTA time, door-to-IVT time, and CSC door-to-EVT time. Functional outcome was assessed using the 90-day modified Rankin Scale (mRS). The Brain magnetic resonance imaging (MRI) after admission was reviewed. A lacune was defined as cytotoxic edema less than 20 mm from the MRI. The definition of medium vessel occlusion (MeVO) remains inconsistent across previous studies^29^. In our study, occlusion of the middle cerebral artery (MCA) M3/M4, anterior cerebral artery (ACA) A2/A3, and posterior cerebral artery (PCA) P2/P3 segments was classified as distal MeVO, while occlusion of the MCA M2, ACA A1, and PCA P1 segments was classified as proximal MeVO, given the uncertain role of EVT for these vessels. The judgement of large core was decided by the interventionalist based on NCCT or CT perfusion (CTP).

### Definition of Futile Transfer

Transfers were defined as “futile” if the patient was transferred for potential EVT but ultimately did not undergo the procedure. Futile transfers were further stratified into three categories.

The first, preventable futile transfers, comprised cases in which pre-transfer vascular imaging could have identified the absence of an actionable target, rendering the transfer unnecessary—specifically: (1) stroke mimics; (2) lacunar strokes confirmed on MRI; (3) TIA without evidence of acute infarction; and (4) no demonstrable large vessel occlusion on CTA, including distal MeVO or scattered emboli.

The second, unpreventable physiological futility, comprised cases in which the transfer decision was appropriate given information available at the PSC, but EVT was ultimately precluded by dynamic biological events during or after transport: (1) spontaneous or post-thrombolytic recanalization, defined as clinical rapid improvement—characterized by an NIHSS score of less than 6 or a decrease of more than 4 points upon CSC arrival—in the context of confirmed ischemic stroke on MRI and thrombus migration to a distal vessel segment on CTA; and (2) post-thrombolytic hemorrhage precluding further intervention.

The third, gray zone, encompassed cases in which the transfer decision was clinically reasonable, but EVT was deferred following re-evaluation at the CSC: (1) large established infarct cores; (2) proximal MeVO; (3) ICAS and CTO; (4) arterial dissection; and (5) socioeconomic or patient-related factors precluding intervention. Gray zone cases do not uniformly represent inappropriate transfers—particularly for large infarct cores, transfer to a CSC may be warranted for higher-level neurological care regardless of EVT eligibility, and the threshold for intervention in this population evolved substantially during the study period. The characterization of gray zone composition is therefore intended to identify targets for system-level improvement, not to render retrospective judgment on individual transfer decisions.

### Statistical Analysis

Patients were stratified according to the timing of CTA acquisition: those undergoing CTA at the PSC before transfer (pre-transfer CTA group) and those undergoing only NCCT at the PSC, with CTA deferred until arrival at the CSC (post-transfer CTA group). Continuous variables are presented as medians with interquartile ranges (IQRs), and categorical variables as frequencies and percentages. Group comparisons were performed using the Mann–Whitney U test for continuous variables and the Chi-square test or Fisher’s exact test for categorical variables. To identify independent predictors of futile transfer, both univariable and multivariable logistic regression analyses were performed. Candidate variables included age, sex, baseline NIHSS score, onset-to-PSC arrival time, and CTA location (Pre-transfer versus post-transfer). Variables were entered into the multivariable model simultaneously. Results are reported as odds ratios (ORs) with 95% confidence intervals (CIs). A 2-sided P-value <.05 was considered statistically significant. All statistical analyses were conducted using IBM SPSS Statistics for Windows, Version 27.0 (IBM Corp., Armonk, NY, USA).

## Result

### Study Population and Baseline Characteristics

A total of 314 patients transferred for potential EVT were included in the analysis after excluding those did not undergo CTA (n=23), had in-hospital stroke onsets or developed LVO symptoms during ED stay in PSC (n=22); (3) was not actually transferred (n=9); or (4) had duplicated records (n=1) (Supplementary Figure S2). Based on the location of vascular imaging, 66 patients (21%) underwent CTA at the PSC (pre-transfer CTA group), while 248 patients (79%) received CTA after arrival at the CSC (post-transfer CTA group) (Table 1). Baseline demographic and clinical characteristics were comparable between the two groups, including age (73 [62–82] vs. 74 [65–82] years; P=0.469), sex (57.6% vs. 56.0% male; P=0.934), and baseline NIHSS score (17 [12–22] vs. 19 [12–24]; P=0.210). The rates of pre-transfer IVT administration were also similar between groups (36.4% vs. 42.7%; P=0.427). The EVT rate differed markedly between groups: 72.7% in the pre-transfer CTA group versus 33.9% in the post-transfer CTA group (P<0.001). Conversely, the futile transfer rate was substantially higher in the post-transfer CTA group (66.1% vs. 27.3%; P<0.001). Overall, 182 of 314 transferred patients (58.0%) did not undergo EVT and were classified as futile transfers (Table 1).

**Table 1.** Baseline Characteristics, Workflow Time Metrics, and Clinical Outcomes of Transferred Patients Stratified by CTA timing.

|  | All | Pre-transfer CTA | Post-transfer CTA | P value |
| --- | --- | --- | --- | --- |
| n | 314 | 66 | 248 |  |
| Age | 73 [64-82] | 73 [62-82] | 74 [65-82] | 0.721 |
| Sex (man, %) | 178 (56.5%) | 38 [57.6%] | 139 (56%) | 0.824 |
| NIHSS | 18 [12-23] | 17 [11.75-22] | 19 [12-24] | 0.311 |
| IVT (%) | 131 (41.6%) | 24 (36.4%) | 106 (42.7%) | 0.350 |
| EVT (%) | 132 (42%) | 48 (72.7%) | 84 (33.9%) | <0.001* |
| Patient receiving EVT |  |  |  |  |
| n | 132 | 48 | 84 |  |
| Age | 73 [63-80] | 73 [62-82.75] | 73 [63.25-80] | 0.987 |
| NIHSS | 18 [12-23] | 17 [11.25-21.75] | 19 [13.25-23.75] | 0.521 |
| IVT (%) | 58 (43.9%) | 17 (35.4%) | 41 (48.8%) | 0.136 |
| O-to-PSC DI time | 79 [44-271.25] | 67.5 [38.5-314] | 83 [44.25-222.5] | 0.587 |
| PSC DIDO time | 99.5 [80.25-149.25] | 151 [102.25-190] | 86 [71.5-106.75] | <0.001* |
| Transfer time | 24 [16.25-31.75] | 30 [25.25-42.25] | 20 [15-27.75] | <0.001* |
| O-to-CSC DI time | 249.5 [178-408] | 289 [220-489] | 212 [163.25-357.75] | 0.007* |
| DI-to-CTA time | 30 [20-52.75] | 54.5 [37-84.75] | 23.5 [18.25-32.75] | <0.001* |
| O-to-CTA time | 228 [153-380.75] | 149.5 [105-379.5] | 239.5 [195.25-385.5] | 0.439 |
| CSC DI-to-EVT | 102.5 [87.25-135.5] | 87.5 [69-113.75] | 109 [96-148] | 0.047 |
| O-to-EVT time | 358.5 [299.25-536.5] | 426 [305-559] | 348 [292.5-483.5] | 0.366 |
| 3-month mRS 0-2 | 42 (31.8%) | 13 (27%) | 29 (34.5%) | 0.521 |

**Footnotes:** Data are presented as n (%) for categorical variables and median [interquartile range] for continuous variables. **Abbreviations:** CTA, Computed Tomography Angiography; DI, Door-in; DIDO, Door-In-Door-Out; EVT, Endovascular Thrombectomy; IVT, intravenous thrombolysis; NIHSS, National Institutes of Health Stroke Scale; mRS, modified Rankin Scale; O, onset. P-values were calculated using the Mann-Whitney U test for continuous variables and Pearson X<sup>2</sup> test or Fisher's exact test for categorical variables.

**Table 2.** Causes of Futile Transfer Stratified by CTA timing.

|  | **Pre-transfer CTA**<br>**(n=18)** | **Post-transfer CTA**<br>**(n=164)** |
| --- | --- | --- |
| **Preventable futile transfer** | **2 (11.1%)** | **66 (40.2%)** |
| Stroke mimic | 0 (0.0%) | 26 (15.9%) |
| Lacunar infarction | 0 (0.0%) | 15 (9.1%) |
| Transient ischemic attack | 0 (0.0%) | 5 (3.0%) |
| Distal MeVO (M3/M4, A2, P2) | 2 (11.1%) | 20 (12.2%) |
| **Unpreventable physiological futility** | **1 (5.6%)** | **14 (8.5%)** |
| Post-IVT hemorrhage | 0 (0.0%) | 2 (1.2%) |
| Spontaneous or post-IVT<br>recanalization | 1 (5.6%) | 12 (7.3%) |
| **Gray zone** | **15 (83.3%)** | **78 (47.6%)** |
| Large core infarction | 3 (16.7%) | 32 (19.5%) |
| ICAS/CTO | 5 (27.8%) | 32 (19.5%) |
| Proximal MeVO (M2, A1, P1) | 2 (11.1%) | 10 (6.1%) |
| Arterial dissection | 2 (11.1%) | 1 (0.6%) |
| Socioeconomic factors (patient<br>condition, family preference,<br>financial issue) | 3 (16.7%) | 3 (1.8%) |
| **Missing data** | **0 (0.0%)** | **6 (3.7%)** |

**Abbreviations:** CTA, Computed Tomography Angiography; CTO, chronic total occlusion; ICAS, intracranial atherosclerotic stenosis; IVT, intravenous thrombolysis; MeVO, Medium Vessel Occlusion.

### Workflow Time Metrics and the Time Trade-off

Pre-transfer CTA was associated with a significant prolongation of PSC DIDO time compared to the post-transfer CTA group, both in the overall cohort (140 [103–181] vs. 88 [76–109] min; P<0.001) (Supplementary Table S1) and in the EVT subgroup (151 [103–188] vs. 86 [72–106] min; P<0.001) (Table 1). Correspondingly, onset-to-CSC arrival time was longer in the pre-transfer CTA group (325 [220–448] vs. 246 [164–415] min; P=0.007). Notably, the onset-to-CTA time was shorter in the pre-transfer CTA group (216 [105–362] vs. 279 [198–450] min; P<0.001), reflecting the earlier acquisition of vascular imaging at the referring center (Supplementary Table S1).

Among patients who underwent EVT, pre-transfer CTA was associated with a significantly shorter CSC door-to-EVT time (88 [71–113] vs. 109 [96–148] min; P<0.001), consistent with reduced need for repeat vascular imaging at the CSC. However, onset-to-EVT time showed a numerical trend toward longer delays in the pre-transfer CTA group (426 [305–553] vs. 348 [294–482] min; P=0.106), though this difference did not reach statistical significance. The rate of favorable functional outcome (mRS 0–2) was numerically lower in the pre-transfer CTA group (27% vs. 34.5%; P=0.521), though this difference also did not reach statistical significance (Table 1).

### Analysis of Futile Transfers

Among the 164 futile transfers in the post-transfer CTA group, 66 cases (40.2%) were classified as preventable—comprising stroke mimics (n=26, 15.9%), lacunar infarctions (n=15, 9.1%), transient ischemic attacks (n=5, 3.0%), and distal medium vessel occlusions (n=20, 12.2%). These represent transfers that would in principle have been avoidable had pre-transfer vascular imaging been performed. An additional 14 cases (8.5%) were attributed to unpreventable physiological futility, including spontaneous or post-thrombolytic recanalization (n=12, 7.3%) and post-thrombolytic hemorrhage (n=2, 1.2%), reflecting dynamic biological events during transport that could not have been anticipated by earlier imaging. The remaining 78 cases (47.6%) fell within the gray zone, with large established infarct cores (n=32, 19.5%) and ICAS/CTO (n=32, 19.5%) constituting the dominant causes, followed by proximal MeVO (n=10, 6.1%), socioeconomic or patient-related factors (n=3, 1.8%), and arterial dissection (n=1, 0.6%). Six cases (3.7%) were unclassifiable due to incomplete records.

In contrast, among the 18 futile transfers in the pre-transfer CTA group, the distribution was markedly different. Only 2 cases (11.1%) were classified as preventable, both involving distal vessel occlusions identified on pre-transfer CTA that were not acted upon at the PSC level, likely reflecting imaging misinterpretation or conservative clinical judgment. One case (5.6%) represented unpreventable physiological futility due to recanalization during transport. The vast majority—15 cases (83.3%)—resided within the gray zone, predominantly driven by ICAS/CTO (n=5, 27.8%), large infarct cores (n=3, 16.7%), socioeconomic factors (n=3, 16.7%), arterial dissection (n=2, 11.1%), and proximal MeVO (n=2, 11.1%).

### Predictors of Futile Transfer

In the univariable analysis, onset-to-PSC arrival time (OR 1.001; 95% CI 1.000–1.002; P=0.014) and post-transfer CTA (OR 5.206; 95% CI 2.851–9.507; P<0.001) were significantly associated with futile transfer, whereas age, sex, and baseline NIHSS were not. In the multivariable model adjusting for these covariates, post-transfer CTA remained the strongest independent predictor of futile transfer (adjusted OR 5.211; 95% CI 2.830–9.595; P<0.001). A longer onset-to-PSC arrival time also independently predicted futility (adjusted OR 1.001; 95% CI 1.000–1.002; P=0.025); although the per-minute effect is modest, its significance across the observed range of arrival times reflects the cumulative time-dependent attrition of EVT eligibility. Notably, patient-level characteristics—including age, sex, and stroke severity—were not independent predictors of futile transfer, suggesting that futility is driven primarily by system-level factors rather than inherent patient characteristics (Table 3).

**Table 3.** Univariable and Multivariable Logistic Regression Analysis of Independent Predictors for Futile Interhospital Transfer.

| Variable | Univariable OR<br>(95% CI) | P | Adjusted OR<br>(95% CI) | P |
| --- | --- | --- | --- | --- |
| Age | 1.009 (0.992–1.027) | 0.281 | 1.010 (0.990–1.030) | 0.328 |
| Sex (Male) | 1.032 (0.657–1.622) | 0.891 | 0.970 (0.584–1.611) | 0.907 |
| Baseline NIHSS | 0.999 (0.970–1.029) | 0.954 | 0.992 (0.960–1.026) | 0.642 |
| Onset-to-PSC Arrival Time (min) | 1.001 (1.000–1.002) | **0.014** | 1.001 (1.000–1.002) | **0.025** |
| Post-transfer CTA (Ref: pre-transfer CTA) | 5.206 (2.851–9.507) | **<0.001** | 5.211 (2.830–9.595) | **<0.001** |

**Footnotes:** The multivariable model was adjusted for age, sex, baseline NIHSS, and onset-to-PSC arrival time. Futile transfer (outcome = 1) is defined as transfer without subsequent endovascular thrombectomy. CTA Location compares patients imaged at the CSC (coded as 1) against those imaged at the PSC (reference, coded as 0). An OR > 1 indicates increased risk of futile transfer. Bold P values indicate statistical significance ( $P < 0.05$ ).
**Abbreviations:** OR, Odds Ratio; CI, Confidence Interval; CTA, Computed Tomography Angiography; NIHSS, National Institutes of Health Stroke Scale.

## Discussion

Our study highlights a central dilemma in the triage of acute ischemic stroke within a distributed stroke network: the trade-off between workflow speed and diagnostic precision. Deferring CTA to the CSC shortened the DIDO time at the PSC and showed a trend toward faster onset-to-EVT time, though this difference was not statistically significant (348 vs. 426 min; P=0.106). Performing CTA at the PSC, by contrast, greatly reduced futile transfers and improved patient selection, but came at the cost of significantly longer PSC processing times. Of note, the pre-transfer CTA group also showed a numerical trend toward lower rates of favorable functional outcome (mRS 0–2: 27% vs. 34.5%; P=0.521) in EVT patients. Although this difference did not reach statistical significance, likely reflecting the limited sample size of EVT-treated patients, these trends warrant careful attention. Importantly, our findings show that this trade-off cannot be captured by time metrics alone—the reasons behind futile transfers were fundamentally different between the two groups, and this difference has important implications for how we design and evaluate stroke transfer systems.

In the post-transfer CTA group, nearly two-thirds of patients did not receive EVT, placing a heavy burden on CSC resources. Among these futile transfers, 40.2% were caused by preventable reasons, including stroke mimics, lacunar infarctions, TIA, and distal MeVO, that could have been identified by pre-transfer CTA. These findings are consistent with the RACECAT post hoc analysis^30^, which also found that vascular imaging at the local stroke center reduced unnecessary transfers and shortened door-to-puncture time at the thrombectomy-capable center. However, a key difference emerged: RACECAT reported no significant prolongation of DIDO time with vascular imaging (78 vs. 76 min; P=0.6), whereas our data showed a much larger delay (140 vs. 88 min; P<0.001). Notably, even patients without vascular imaging in RACECAT achieved a median DIDO of only 76 minutes, faster than either group in our cohort. This suggests that the gap reflects not simply the presence or absence of CTA, but a broader difference in system-level standardization.

Our data suggest that the delay is attributable to two modifiable processes at PSC: the time from arrival to CTA acquisition, and the time from CTA completion to transfer decision. Regarding the first bottleneck, while universal CTA strategies have been advocated in well-resourced settings^31^, such an approach is not feasible in networks similar to TCSN, where CTA availability and technician coverage are inconsistent. A more practical alternative is a selective CTA protocol triggered by validated high-risk clinical criteria, such as a positive Rapid Arterial Occlusion Evaluation (RACE) score^32^ or an elevated Los Angeles Motor Scale (LAMS) score^33^, to identify patients with a high probability of LVO before imaging. This approach concentrates limited CTA resources on patients most likely to benefit, avoids unnecessary delays for lower-risk cases, and provides a structured decision framework that reduces reliance on individual physician judgment in time-pressured settings. Regarding the second bottleneck, AI-based LVO detection software could provide rapid preliminary reads, reducing dependence on after-hours radiology reporting^34, 35^. On the CSC side, our data confirm that pre-transfer CTA significantly shortened CSC door-to-EVT time (88 vs. 109 min; P<0.001), consistent with the ability to bypass repeat imaging and proceed directly to the angio-suite. Systematic pre-notification of the interventional team at the time of PSC transfer decision, combined with protocol-driven angio-suite activation, could further reduce this interval and help offset upstream delays^36^. Nevertheless, even with optimized workflows, a substantial proportion of futile transfers will persist, driven not by imaging delays but by the complexity of the clinical decisions that follow.

Indeed, in the pre-transfer CTA group, preventable futile transfers were nearly eliminated, confirming the effectiveness of pre-transfer CTA as a triage filter. However, gray zone transfers remained the dominant category in both groups, suggesting that pre-transfer CTA alone is insufficient to resolve all sources of transfer inefficiency. Physiological futility, comprising spontaneous or post-IVT recanalization and hemorrhage, accounted for less than 10% of futile transfers in both groups, a proportion notably lower than the recanalization rates of approximately 30% in IVT-treated patients and 10% in untreated patients reported in prior Western cohorts ^37–40^. While some recanalized patients may have been captured within the distal MeVO or no-LVO categories, the overall proportion remained low.

The gray zone represented the largest unresolved category in both groups, dominated by three distinct subgroups—ICAS/CTO-related lesions, large infarct cores, and medium vessel occlusions. Importantly, many of these transfers should not be considered truly futile in the conventional sense. Patients with complex vascular lesions, large infarct cores, or challenging MeVO cases are often transferred not solely for EVT, but because CSCs offer resources and expertise that PSCs cannot provide, including neurosurgical backup, intensive neuromonitoring, and multidisciplinary decision-making. In this context, transfer serves a broader clinical purpose beyond procedural intervention, and reducing these transfers indiscriminately risks denying patients access to the level of care their condition demands.

Among these three subgroups, ICAS/CTO-related lesions were the most prevalent in both groups, reflecting the well-recognized high prevalence of ICAS in East Asian populations^19, 20^, a vascular epidemiological feature largely absent from Western futile transfer literature. Single-phase CTA, the most commonly available modality at PSCs, is often insufficient to reliably distinguish acute thrombus from underlying chronic stenosis, and PSC physicians without immediate access to neuroradiologists or interventionalists may lack the confidence to act on ambiguous findings. Addressing this subgroup will require strategies beyond pre-transfer CTA, including multiphase CTA or CT perfusion^41–44^ and stronger real-time communication between PSC clinicians and CSC specialists. MeVO and large core infarction represent a different but equally complex challenge, where treatment decisions remain highly operator-dependent and continue to evolve. For MeVO, the role of EVT remains uncertain and is typically decided on a case-by-case basis^5^. For large core infarcts, the recent consolidation of EVT evidence has fundamentally expanded the therapeutic landscape^3, 4^, and the proportion of such patients transferred to CSCs is likely to increase as this evidence is adopted into routine practice.

The generalizability of our findings regarding futile transfer may extend to other regional stroke networks. However, the impact of CTA location on transfer efficiency should be interpreted in the context of local healthcare geography. In our network, the high density of stroke centers and relatively short inter-hospital distances result in inherently brief transfer times; consequently, imaging-related delays at the PSC may represent a proportionally greater time penalty. In contrast, in geographically dispersed regions such as North America, Europe, or Australia, where a single futile transfer may require hours of transport and substantial resource utilization, the balance may shift. In such settings, the potential benefit of pre-transfer CTA in reducing unnecessary transfers may outweigh the additional upstream delay.

In dense networks such as ours, however, the margin for delay is narrower, and the emphasis should be placed on optimizing the timeliness and quality of CTA acquisition and interpretation at the PSC, rather than uniformly advocating its routine use. Taken together, these findings suggest that the value of pre-transfer CTA is context-dependent, and that regional stroke systems should tailor triage strategies according to their geographic, infrastructural, and workflow characteristics.

Several limitations of this study warrant acknowledgment. First, as a retrospective observational study, group assignment was not randomized, and residual confounding from unmeasured variables—such as PSC physician experience, time of day, and individual institutional protocols—cannot be excluded. Second, PSC imaging data were not systematically available, limiting our ability to determine whether large infarct cores were already established prior to transfer. Third, 90-day mRS data were collected only for EVT-treated patients, preventing a comprehensive assessment of functional outcomes across the entire transferred cohort. Finally, our findings reflect the specific characteristics of a dense hospital network and should be interpreted with caution before being generalized to stroke systems with different geographic, infrastructural, or workflow profiles.

## Conclusion

In this real-world analysis of a regional stroke transfer network, pre-transfer CTA at the PSC was associated with a substantially lower rate of futile transfer and a higher EVT yield, supporting its role as a precision triage tool in the DS model. However, this benefit came at the cost of significantly prolonged PSC processing times, and numerical trends toward longer onset-to-EVT time and worse functional outcomes further suggest the importance of minimizing imaging-related delays. Beyond preventable transfers, the gray zone, dominated by ICAS/CTO-related lesions, large infarct cores, and MeVO, represents the next frontier of transfer inefficiency, one that cannot be resolved by imaging alone and requires enhanced real-time collaboration between PSCs and CSCs. Ultimately, the value of pre-transfer CTA is context-dependent: regional stroke systems should calibrate their triage strategies according to their own geographic, infrastructural, and workflow realities, with the shared goal of balancing triage precision and time efficiency to achieve optimal patient outcomes.

**Figure 1.**
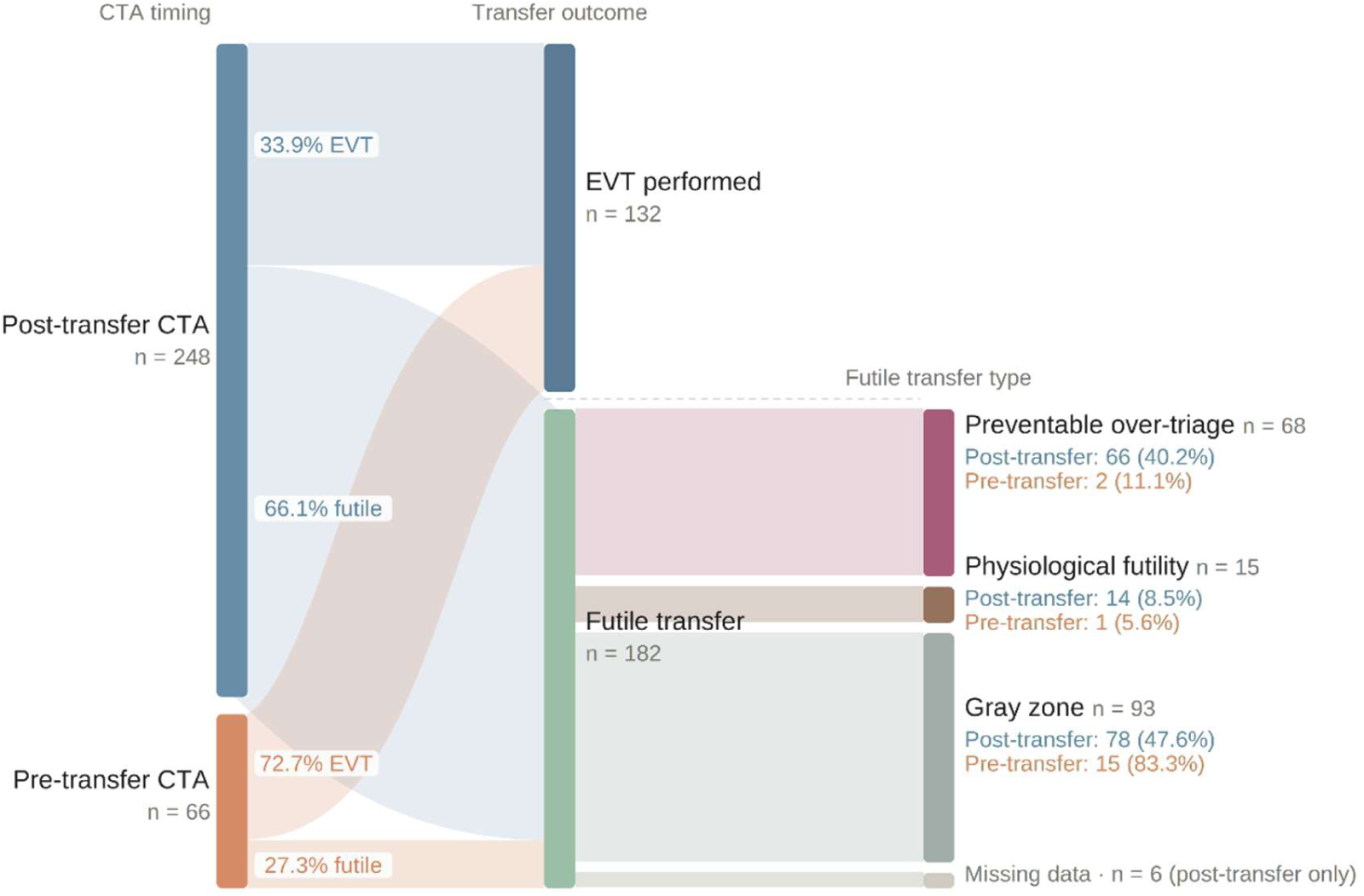
Patient flow and futile transfer classification stratified by CTA timing in an interhospital stroke transfer network. Each node width is proportional to the number of patients. Of 314 patients transferred for endovascular thrombectomy (EVT) evaluation, 248 underwent CTA after arrival at the comprehensive stroke center (post-transfer CTA group) and 66 underwent CTA at the primary stroke center prior to transfer (pre-transfer CTA group). EVT was performed in 84 (33.9%) and 48 (72.7%) patients in the post-transfer and pre-transfer CTA groups, respectively. Among the 182 patients with futile transfer, subtypes were classified into three categories: preventable over-triage, physiological futility, and gray zone. Preventable over-triage accounted for 40.2% of futile transfers in the post-transfer CTA group versus 11.1% in the pre-transfer CTA group, whereas gray zone cases predominated in the pre-transfer CTA group (83.3% vs. 47.6%). **Abbreviations:** CTA, computed tomography angiography; EVT, endovascular thrombectomy.

**Figure 2.**
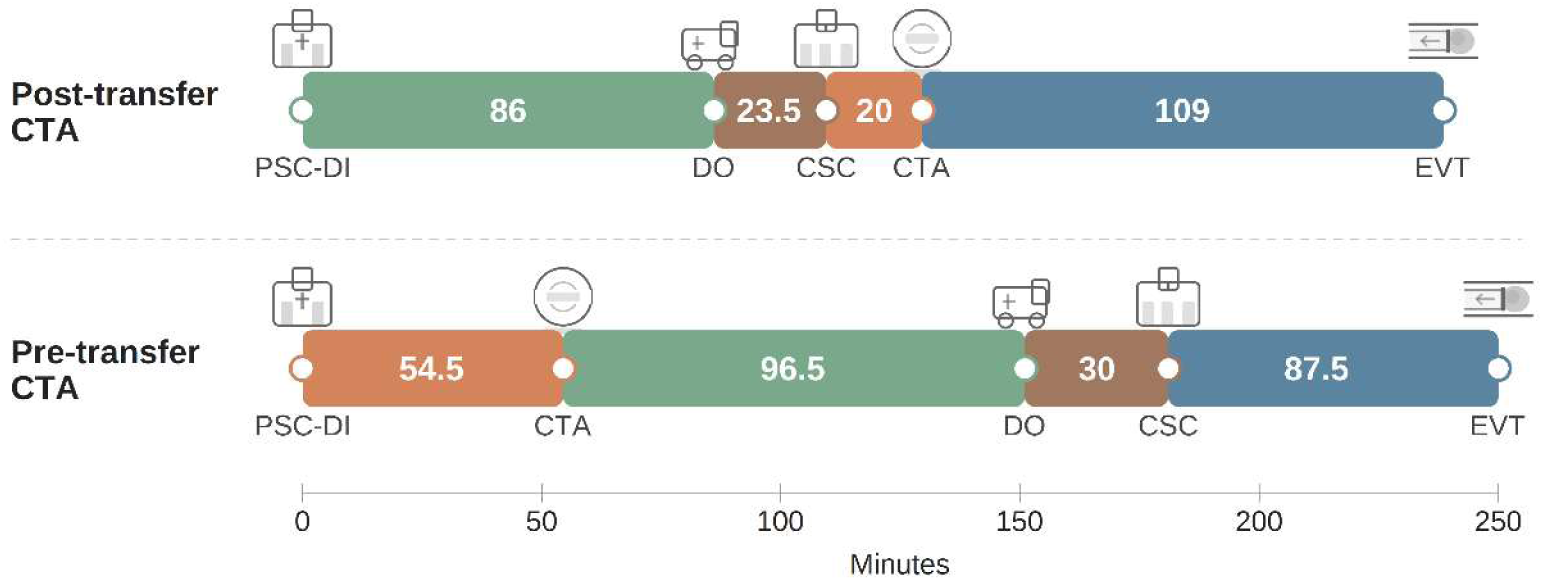
Comparative timeline of median workflow intervals between pre-transfer CTA and post-transfer CTA group. Figure Legend: The timeline compares the median duration (in minutes) of key workflow phases for patients undergoing pre-transfer CTA (bottom row) versus those undergoing post-transfer CTA (top row). While the pre-transfer CTA strategy was associated with a longer median Door-in-Door-out (DIDO) time at the primary hospital (due to image acquisition), it significantly reduced the in-hospital processing time at the receiving CSC. Numbers represent median time in minutes. **Abbreviations:** PSC, Primary Stroke Center; CTA, Computed Tomography Angiography; DI, door-in; DO, door-out; CSC, Comprehensive Stroke Center; EVT, Endovascular Thrombectomy.

**Supplementary Figure S1.**
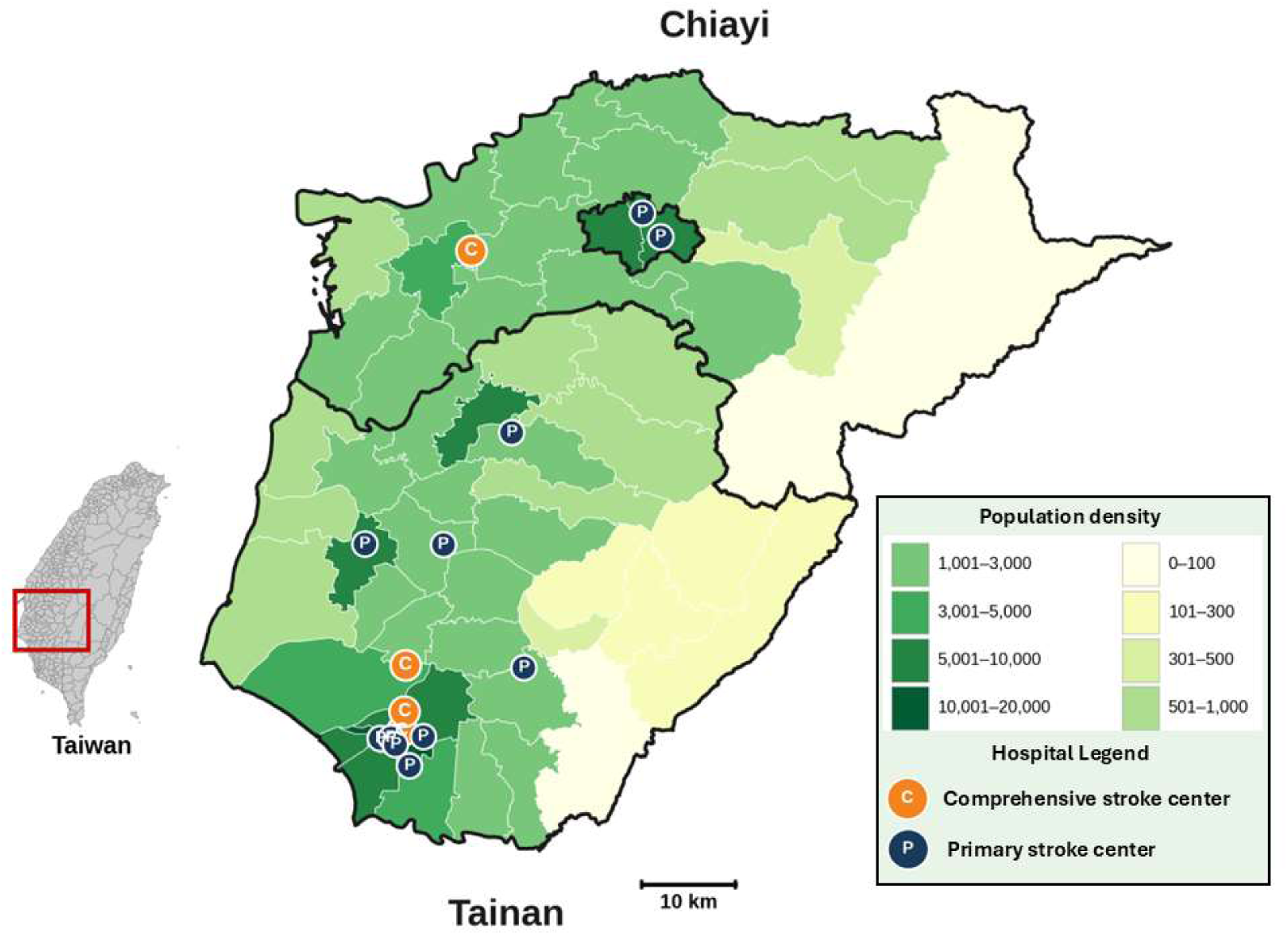
Geographic Distribution of Stroke Centers and Population Density in the Tainan-Chiayi Stroke Network. The map depicts the Tainan-Chiayi region of southern Taiwan (inset), encompassing the administrative districts of Tainan City and Chiayi City and Chiayi County. Township-level population density (persons per km²) is represented by a graduated green color scale, with darker shades indicating higher density. Comprehensive stroke centers (CSCs) and primary stroke centers (PSCs) participating in the Tainan-Chiayi Stroke Network (TCSN) are marked by orange and navy blue icons, respectively. The network comprises 4 CSCs and 11 PSCs distributed across both urban and rural districts, reflecting the regional drip-and-ship transfer system for endovascular thrombectomy (EVT) candidate triage.

**Supplementary Figure S2.**
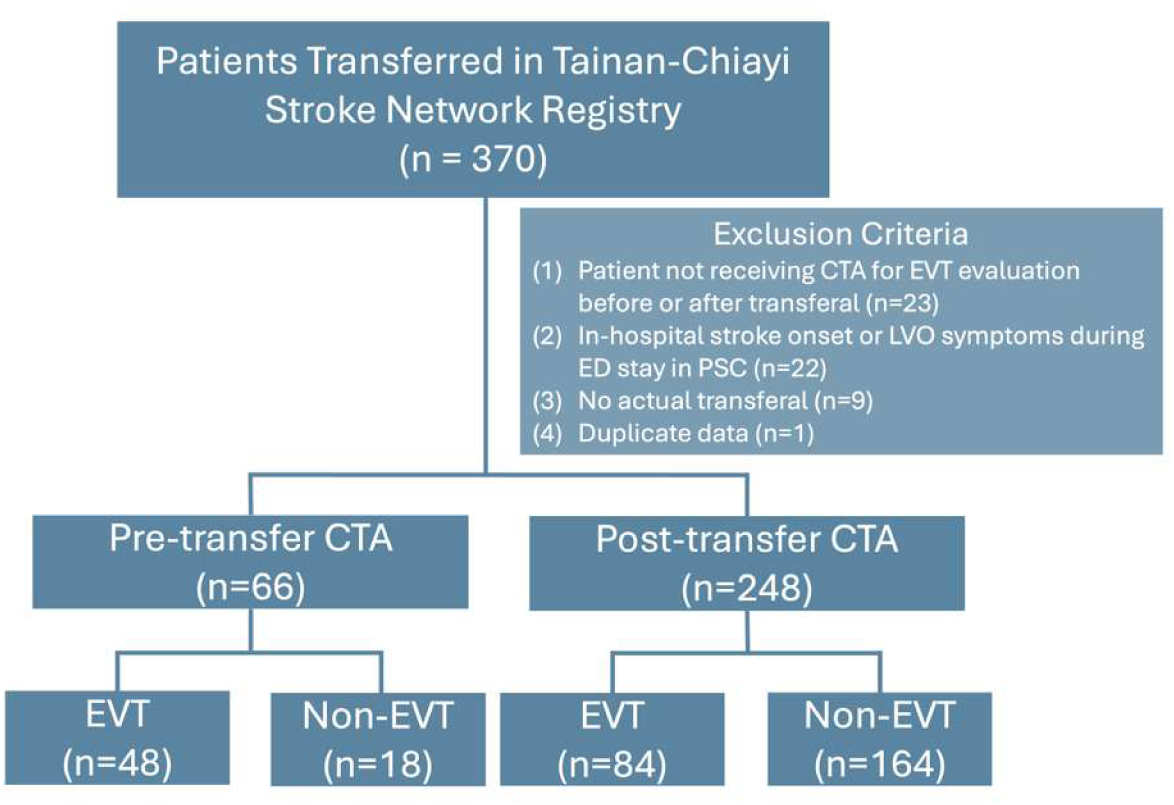
Flowchart of Patient Enrollment from the Tainan-Chiayi Stroke Network Registry. A total of 370 patients transferred within the Tainan-Chiayi Stroke Network (TCSN) were screened for eligibility. Fifty-five patients were excluded: 23 did not receive computed tomography angiography (CTA) for endovascular thrombectomy (EVT) evaluation before or after transfer, 22 had in-hospital stroke onset or developed large vessel occlusion (LVO) symptoms during emergency department stay at the primary stroke center (PSC), 9 had no actual transfer recorded, and 1 had duplicate data. The remaining 314 patients were stratified by CTA timing into two groups: the Pre-transfer CTA group (n=66), in which CTA was performed at the PSC before transfer, and the Post-transfer CTA group (n=248), in which CTA was performed at the comprehensive stroke center (CSC) after transfer. Within each group, patients were further categorized according to whether they ultimately received EVT (Pre-transfer CTA: EVT n=48, non-EVT n=18; Post-transfer CTA: EVT n=84, non-EVT n=164).

**Supplementary Table S1.** Detailed Workflow Time Metrics of Transferred Patients Stratified by CTA Timing.

|  | All | Pre-transfer CTA | Post-transfer CTA | P value |
| --- | --- | --- | --- | --- |
| n | 314 | 66 | 248 |  |
| O-to-PSC DI time | 107.5 [45.75-298.5] | 79.5 [38.75-308.75] | 111 [47-297] | 0.890 |
| PSC DIDO time | 96 [79-130.25] | 140.5 [102.75-182.5] | 88 [76-109] | <0.001* |
| Transfer time | 21 [15-30] | 31.5 [26-49] | 19 [15-26] | <0.001* |
| O-to-CSC DI time | 260 [175.75-431.5] | 325 [219.25-453.75] | 245.5 [163.25-415] | 0.213 |
| O-to-CTA time | 266 [186-437] | 215.5 [103-369] | 279 [197.5-454.75] | 0.143 |
| PSC DI-to-TPA time | 51 [43.5-62.5] | 58.5 [51-72.75] | 51 [42-60] | 0.038* |
| Patient receiving EVT |  |  |  |  |
| N | 132 | 48 | 84 |  |
| O-PSC DI time | 79 [44-271.25] | 67.5 [38.5-314] | 83 [44.25-222.5] | 0.587 |
| PSC DIDO time | 99.5 [80.25-149.25] | 151 [102.25-190] | 86 [71.5-106.75] | <0.001* |
| Transfer time | 24 [16.25-31.75] | 30 [25.25-42.25] | 20 [15-27.75] | <0.001* |
| O-to-CSC DI time | 249.5 [178-408] | 289 [220-489] | 212 [163.25-357.75] | 0.007* |
| O-to-CTA time | 228 [153-380.75] | 149.5 [105-379.5] | 239.5 [195.25-385.5] | 0.439 |
| PSC DI-to-TPA time | 51 [44.75-63.25] | 65 [51.5-75] | 51 [43-57.5] | 0.008* |
| CSC DI-to-EVT | 102.5 [87.25-135.5] | 87.5 [69-113.75] | 109 [96-148] | 0.047* |
| O-to-EVT time | 358.5 [299.25-536.5] | 426 [305-559] | 348 [292.5-483.5] | 0.366 |

**Footnotes:** Data are presented as n (%) for categorical variables and median [interquartile range] for continuous variables. **Abbreviations:** PSC, Primary Stroke Center; CSC, Comprehensive Stroke Center; CTA, Computed Tomography Angiography; DI, Door-in; DIDO, Door-In-Door-Out; EVT, Endovascular Thrombectomy; NIHSS, National Institutes of Health Stroke Scale; O, Onset. Analysis of workflow metrics (CSC Door-to-Puncture, Onset-to-Puncture) and functional outcomes (mRS) includes only patients who underwent EVT. (n=132).
P-values were calculated using the Mann-Whitney U test for continuous variables and Pearson X<sup>2</sup> test or Fisher's exact test for categorical variables.

## Data Availability

The data that support the findings of this study are not publicly available due to patient privacy regulations and institutional data governance policies. De-identified data may be available from the corresponding author upon reasonable request, subject to approval by the institutional review board.

## Acknowledgments

The authors are also grateful to members of the Tainan Stroke Network and Public Health Bureau, Tainan City Government for the support.

Artificial intelligence assistance (Claude, Anthropic) was used for English language editing and figure preparation in this manuscript. The authors assume full responsibility for the accuracy and integrity of all content.

## Sources of Funding

National Cheng Kung University Hospital: NCKUH-11502001, NCKUH-11503023 Ministry of Science and Technology: MOHW 114-2314-B-006-042-MY2; MOHW 114-2622-E-006-041- Both funders do not involve in the study design, the collection/analysis and interpretation of the data; the writing of the report and the decision to submit the paper for publication. National Cheng Kung University Hospital did not influence the results/outcomes of the study despite authors affiliation with the funder.

## Disclosures

The authors declare no conflict of interest.

## Author Contributions

PYT, CMW, and PSS researched literature and conceived the study. PYT, CMW, PSS were involved in protocol development and gaining ethical approval. All authors were involved in patient recruitment and data analysis. PYT wrote the first draft of the manuscript. CMW reviewed and edited the manuscript. All authors approved the final version of the manuscript.

## Ethical approval

Approval for this study was obtained from the Institutional Review Board of National Cheng Kung University Hospital (IRB Approval No. A-ER-109-561; B-ER-114-293; A-ER-115-009).

## Data Availability Statement

The collected data will be available upon reasonable request from the corresponding author.

## Non-standard Abbreviations and Acronyms

AIS: acute ischemic stroke
ACA: anterior cerebral artery
CI: confidence interval
CSC: comprehensive stroke center
CTA: computed tomography angiography
CTO: chronic total occlusion
DIDO: door-in-door-out
DS: drip-and-ship
EVT: endovascular thrombectomy
EVTTS: endovascular thrombectomy transfer system
ICAS: intracranial atherosclerotic stenosis
IVT: intravenous thrombolysis
LAMS: Los Angeles Motor Scale
LVO: large vessel occlusion
MCA: middle cerebral artery
MeVO: medium vessel occlusion
mRS: modified Rankin Scale
MRI: magnetic resonance imaging
MS: mothership
NIHSS: National Institutes of Health Stroke Scale
OR: odds ratio
PCA: posterior cerebral artery
PSC: primary stroke center
TCSN: Tainan-Chiayi Stroke Network
TIA: transient ischemic attack

## Notes

### Competing Interest Statement

The authors have declared no competing interest.

### Clinical Trial

Not applicable

### Funding Statement

This work was supported by National Cheng Kung University Hospital (grant numbers NCKUH-11502001 and NCKUH-11503023) and the Ministry of Health and Welfare (grant numbers MOHW 114-2314-B-006-042-MY2 and MOHW 114-2622-E-006-041). The funders had no role in study design, data collection, analysis, or interpretation, manuscript preparation, or the decision to submit for publication. Although the authors are affiliated with National Cheng Kung University Hospital, the institution did not influence the results or outcomes of this study. No payment or services from any third party were received for any aspect of the submitted work.

